# Safety and superior immunogenicity of heterologous boosting with an RBD-based SARS-CoV-2 mRNA vaccine in Chinese adults

**DOI:** 10.1101/2022.05.30.22275753

**Authors:** Xiaoqiang Liu, Yuhua Li, Zhongfang Wang, Shouchun Cao, Weijin Huang, Lin Yuan, Yi-Jiao Huang, Yan Zheng, Jingjing Chen, Bo Ying, Zuoyun Xiang, Jin Shi, Jincun Zhao, Zhen Huang, Cheng-Feng Qin

## Abstract

Homologous and heterologous booster with COVID-19 mRNA vaccines represent the most effective strategy to prevent the ongoing Omicron pandemic. The additional protection from these prototype SARS-CoV-2 S-targeting vaccine was attributed to the increased RBD-specific memory B cells with expanded potency and breadth. Herein, we show the safety and immunogenicity of heterologous boosting with the RBD-targeting mRNA vaccine AWcorna (also term ARCoV) in Chinese adults who have received two doses inactivated vaccine. The superiority over inactivated vaccine in neutralization antibodies, as well as the safety profile, support the use of AWcorna as heterologous booster in China.

## Introduction

To May 2022, the COVID-19 pandemic has claimed more than 6.28 million lives, with more than 524 million confirmed cases worldwide. The recent emergence of highly transmissible Omicron variant of severe acute respiratory syndrome coronavirus 2 (SARS-CoV-2) has triggered another major surge in both confirmed cases and deaths^1^. Ten COVID-19 vaccines have been approved by the World Health Organization (WHO) for emergency use, including the two mRNA vaccines, BNT162b2 and mRNA-1273, and two Chinese inactivated vaccines, CoronaVac and BBIBP-CorV. However, the rapid waning of vaccine-induced virus-neutralizing antibody titers and the continuous emergence of variants of concern (VOCs), including Alpha, Beta, Delta and Omicron, have created unprecedented challenges in the eradication of COVID-19 pandemic^2-4^.

Especially, the heavily mutated Omicron variant has been well characterized to escape from most therapeutic monoclonal antibodies, as well as sera from convalescent patients or fully vaccinated individuals^5,6^. Recent data has indicated that neutralizing antibody against Omicron was absent or undetectable in most Chinese populations who received two-dose inactivated vaccines^7^. Interestingly, preliminary data have suggested that a booster dose with mRNA vaccine BNT162b2 showed significant superiority over homologous booster in titers and protection against Omicron^7-9^. However, the two commercial mRNA vaccines, encoding the full S protein of SARS-CoV-2, are not available in mainland China. The “made-in-China” mRNA vaccine candidate AWcorna (originally termed ARCoV), which encodes the Receptor Binding domain (RBD) of SARS-CoV-2 S protein, is being tested in the final stage of multiple-center phase III trials (https://clinicaltrials.gov/ct2/show/NCT04847102). It is of highly priority and urgency to provide evidence to support a better boosting strategy in mainland China for decision maker.

## Results

Herein, we reported the safety and immunogenicity of a third dose of heterologous boosting with AWcorna in Chinese adults who have received two-dose inactivated vaccines. The randomized clinical trial (ChiCTR2100053701) enrolled 300 adults (ages≥18 years). All eligible subjects received 2-dose priming vaccination with the inactivated vaccine, CoronaVac or BBIBP-CorV. At about 6-month post-priming, all subjects were randomly assigned to either the AWcorna (n=200; heterologous) or CoronaVac (n=100; homologous) booster group (Supplementary information, **Fig. S1a**). In the AWcorna group, the median age was 43.0 years (Interquartile rate, IQR: 36.5-49.0), and the CoronaVac group was 40.0 years (IQR: 34.0-48.5) (*P*=0.5165) (Supplementary information, **Table S1**). There were 116 (58%) and 55 (55%) male participants in the AWcorna and CoronaVac groups, respectively (*P*=0.6208). Meanwhile, no significant differences were observed in the Body Mass Index (BMI), vital signs, and comorbid condition between the two groups at the baseline (All *P*>0.05). All subjects completed the enrollment vaccination and three blood examinations consecutively at pre-booster or 0 days, 14±2 days, and 28±2 days post -booster vaccination. Subsequently, the neutralization and IgG antibody titers against wild-type (WT) SARS-CoV-2 and VOCs were assessed at pre-booster, 14- and 28-day post-booster by the standard cytopathic effect (CPE) based assay and ELISA, respectively (Supplementary information, Methods). The WHO standard IgG antibody (NIBSC code 20/136) was used as a reference sample for all serological assays.

As expected, the live virus neutralization titers against WT SARS-CoV-2 were below the detection limit before boosting in all participants from both groups (**Fig. 1a**). Remarkably, AWcorna booster induced a 66.59-fold increase against WT SARS-CoV-2, and the geometric mean titers (GMTs) reached 293.9 and 242.4 at 14 and 28 days post booster, respectively (WHO Reference cut-off 1:139), while the GMTs in CoronaVac booster groups was only 89.1 and 64.3, respectively (**Fig. 1a** and supplementary information, **Table S2)**. Similarly, the neutralization antibody titers against the Delta variant also increased significantly after the third dose booster either with AWcorna and CoronaVac, while the GMTs in Awcorna groups were 5.1- and 6.5-fold higher than that in CoronaVac group at 14 and 28 days post booster (**Fig. 1b** and supplementary information, **Table S2**). In addition, the increasing trends are similar in both 18-59 and ≥60 years old participants (Supplementary information, **Fig. S2a**,**b**).

**Fig.1.**
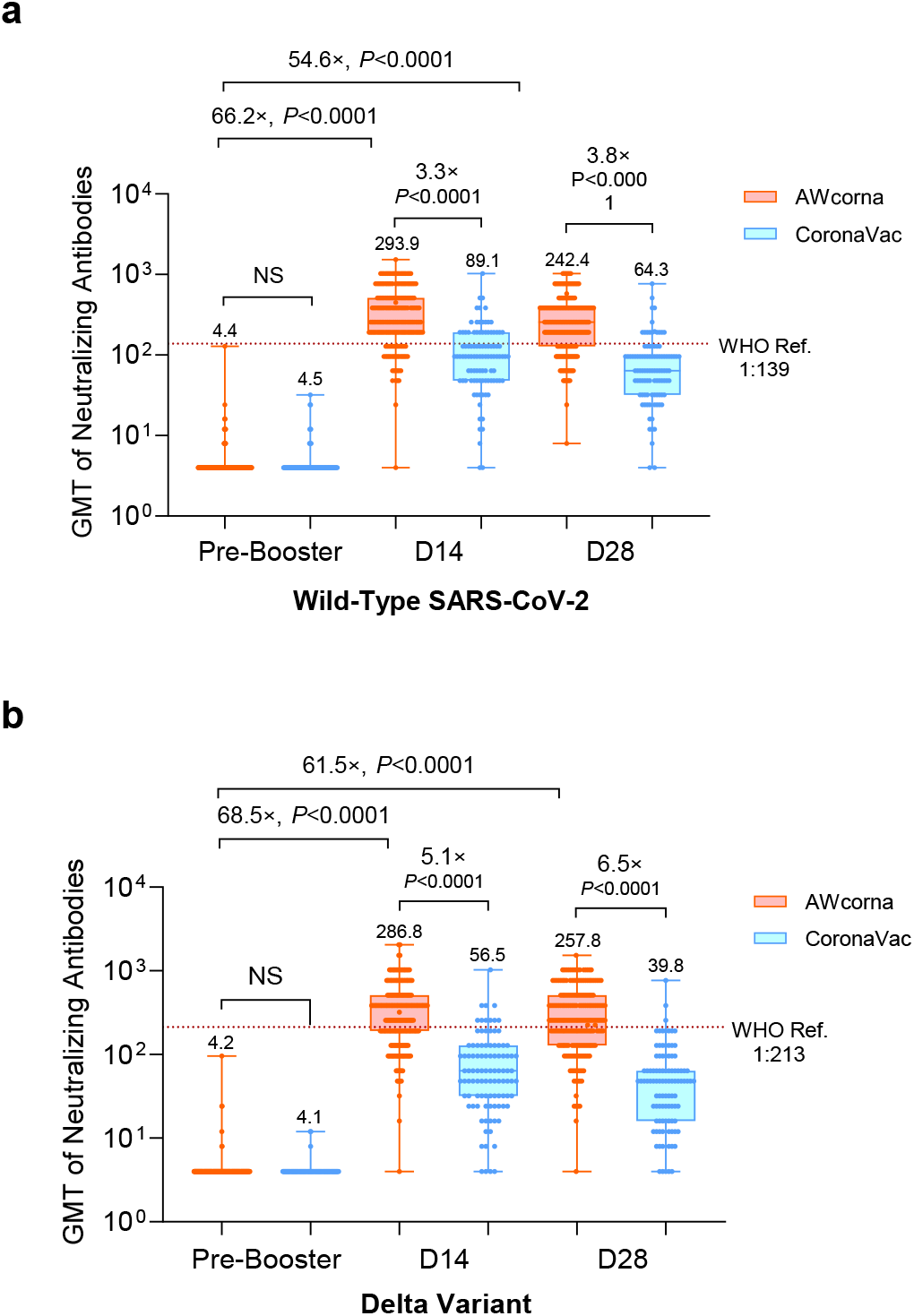
GMT of neutralizing antibodies to live wild-type SARS-CoV-2 or Delta variant. **a** Geometric mean titer (GMT) of neutralizing antibodies to live wild-type (WT) SARS-CoV-2. The WHO reference serum (1,000 IU/ml) was equivalent to a live viral neutralizing antibody titer of 1:139 against WT. **b** GMT of neutralizing antibodies to the Delta variant. The WHO reference was 1:213 against the Delta variant (the dash line in red). Eligible participants primed with 2-does of inactivated vaccine were randomly allocated to AWcorna group (n=200) and CoronaVac group (n=100) to receive a booster dose respectively. GMT data are presented in box-and-whisker plots. The figures above error bars indicate the percentage. *P* values were obtained from comparisons between the two treatment groups using *t*-tests for log-transformed antibody or two-sided χ2 tests for categorical data.

Despite the neutralization antibody titers against the Omicron variant showed significant reduction in comparison with that against the WT virus in both groups, the GMTs against Omicron maintained 28.1 at 28-day after AWcorna booster, while the GMT in the CoronaVac booster group was only 6.4 (**Fig. 2a** and supplementary information, **Table S2**). Most importantly, 83.75% of participants in the AWcorna booster group achieved the 1:8 threshold of neutralization antibody titers against the Omicron relative to that of only 35% of participants in the CoronaVac booster group (**Fig. 2b**; 95% Confidence Interval, CI: 30.82-63.84; *P*<0.0001).

**Fig.2.**
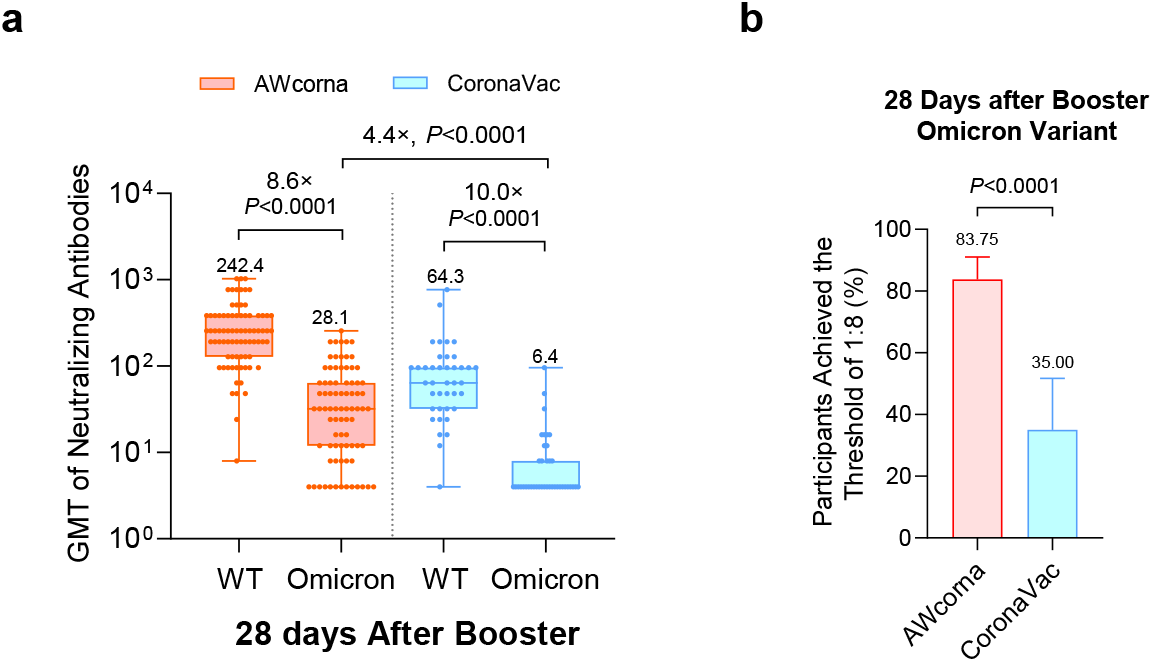
GMT of neutralizing antibodies to Omicron variant. **a** GMT of neutralizing antibodies to the Omicron variant was measured in the subgroup (120 participants with the first 120 subject numbers, 80 from AWcorna and 40 from CoronaVac group). Sera were measured 28 days after booster only, thus no baseline analysis was performed. **b** Seropositive rates (%) of neutralizing antibody to the Omicron variant. GMT data are presented in box-and-whisker plots. The figures above error bars indicate the percentage. *P* values were obtained from comparisons between the two treatment groups using *t*-tests for log-transformed antibody or two-sided χ2 tests for categorical data.

Moreover, the RBD-specific IgG antibodies titers also showed a sharp increasement in both booster groups, and the GMT in AWcorna booster group was 6.8- and 7.1-fold higher than that in the CoronaVac booster group at both 14-day and 28-day time points, respectively (all *P*<0.0001) (**Fig. 3a** and supplementary information, **Table S3)**. Taken together, these results demonstrate that heterologous boosting with AWcorna induces higher neutralization and IgG antibodies against WT, Delta and Omicron variants than homologous booster.

**Fig.3.**
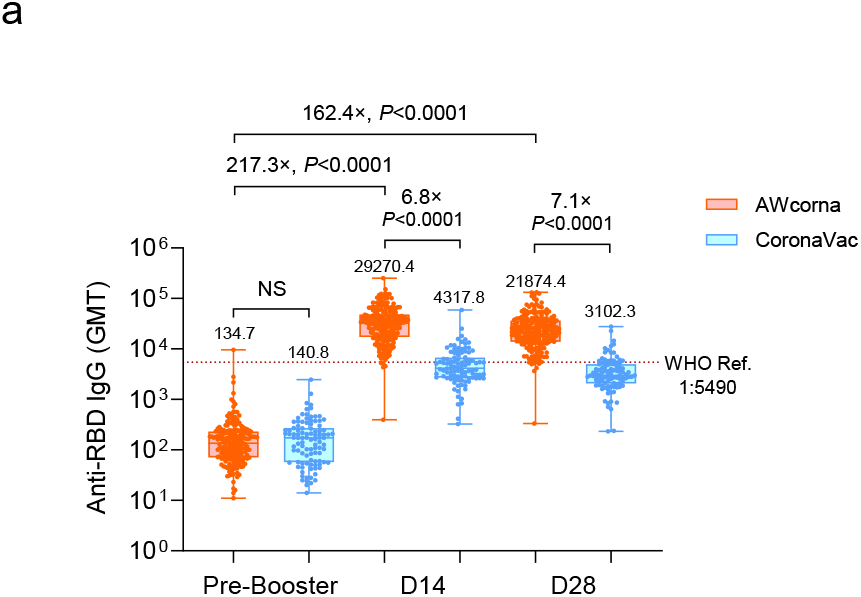
GMT of anti-RBD IgG antibodies to WT SARS-CoV-2. **a** GMTs of anti-RBD IgG antibodies to WT SARS-CoV-2. The WHO reference (1,000 binding antibody unit (BAU)/ml in serum) is equivalent to an RBD-specific IgG ELISA antibody titer of 1:5490. The cutoff value for the response was 1:8 for live virus neutralizing antibody and 1:10 for anti-RBD IgG. GMT data are presented in box-and-whisker plots. The figures above error bars indicate the percentage. *P* values were obtained from comparisons between the two treatment groups using *t*-tests for log-transformed antibody or two-sided χ2 tests for categorical data.

Additionally, we observed the safety profile of the booster dose of AWcorna. Solicited local and systemic adverse events (AEs) were recorded within 30 mins and in a window of 0-14 days, and unsolicited AEs were documented within 0-28 days post-booster vaccination (Supplementary information, **Table S4**). For both vaccines, pain at the injection site is the most reported local AE (incidence rate, IR:17% in AWcorna vs. 2% in CoronaVac; *P*<0.0001), mostly at the Grade 1 level (**Fig. 4a**). Fever was the most common systemic AE (IR: 33.5%), followed by headache (IR: 26.0%) and muscle aches (IR: 7.5%) in AWcorna group (**Fig. 4b**). A total of 8 subjects reported grade 3 fever (IR:4%) among the 200 participants in AWcorna group. For the CoronaVac group, headache represented as the most frequent systemic AE (IR: 7.0%), followed by fever (IR: 4.0%). No serious adverse events (SAE) were reported in both groups.

**Fig.4.**
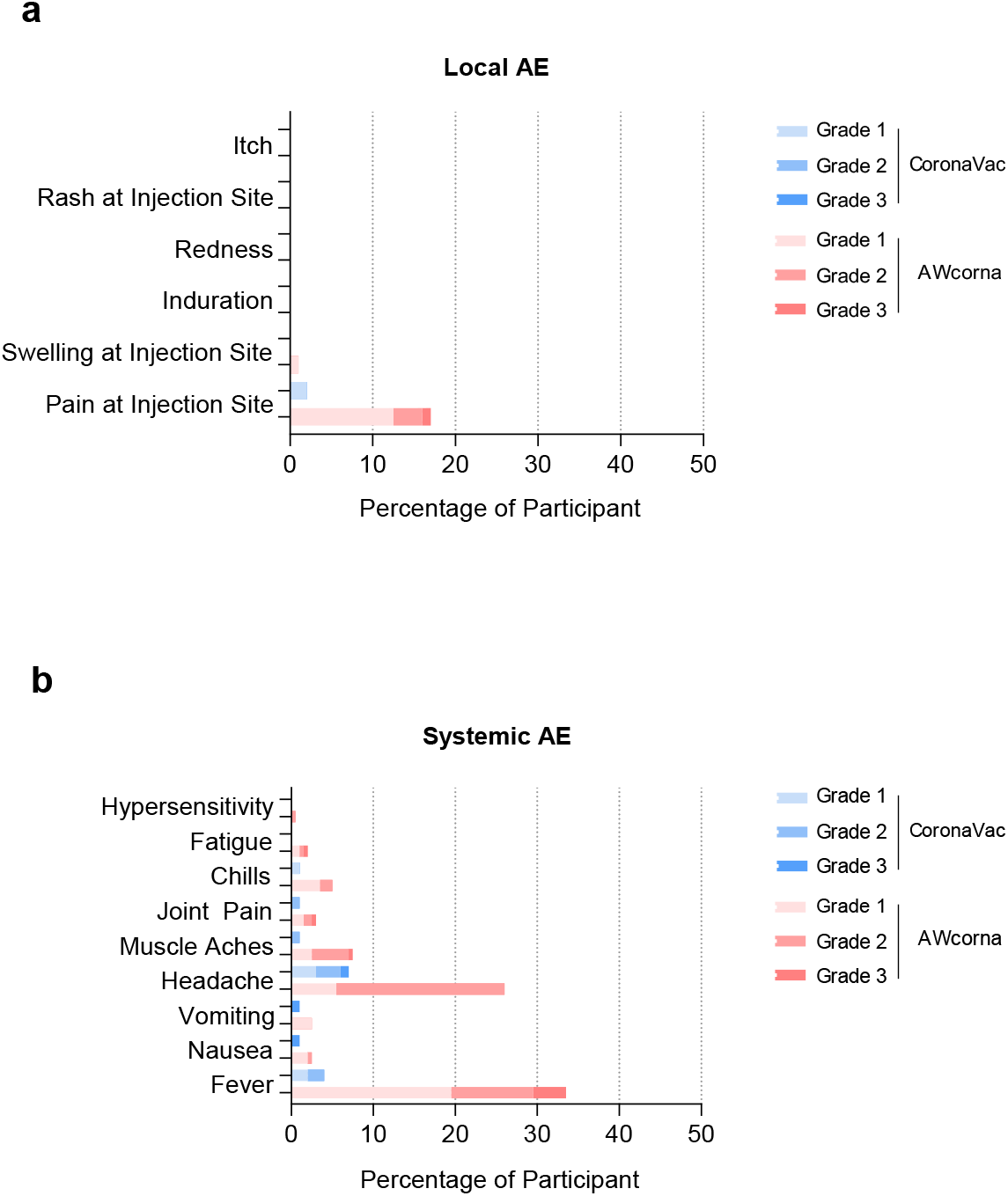
Safety of Heterologous boosting with AWcorna in Chinese adults. **a** The percentage of participants with local adverse events (AEs). **b** The percentage of participants with systemic AEs. These AEs were monitored in the 14 days window after the administration of the booster.

## Discussion

Collectively, our present study clearly demonstrated that a 3^rd^ dose of heterologous boosting with AWcorna was safe and potential protective against the circulating Delta and Omicron variants. Compared with phase 1 trial, the total IRs of local and systemic AEs for AWcorna booster showed significant improvement^10^, especially the IR of grade 3 fever reduced to 4% (Supplementary information, **Table S4**), which was comparable to the other two approved mRNA vaccines^11^. The phase 1 trial of AWcorna only included 20 adults aged 18-59 (15 μg group), while our present cohort enrolled 200 participants, including 10 subjects aged over 60. The expansion of sample size and improvement in vaccine manufacturing technologies contributed to the improved safety profile observed in our present study. The ongoing international phase 3 trials with 28,000 participants will provide more meaningful data about the safety profile of AWcorna.

Previously, we have demonstrated that homologous boosting with AWcorna readily induced high neutralization antibodies against WT and Omicron variant in mice^12^, our present data in human further supported heterologous booster with this China-made mRNA vaccine AWcorna in Chinese populations. Many cities in China are under the attack of Delta and Omicron variants, while few or no neuralization antibodies against Omicron was detected in most Chinese populations^7,13^, a third dose of booster has been recommended by the WHO and National Health Commission of China. Of all COVID-19 vaccines generated from different technology platform, mRNA vaccine represents as the most reasonable choice as either homologous or heterologous booster. The neutralization titers against Delta and Omicron variants in AWcorna booster group were 6.5-fold and 4.4-fold higher than that in CoronaVac booster group, respectively (**Fig. 1b, c**), and the AWcorna booster induced the seroconversion of Omicron neutralization in over 83% individuals (**Fig. 1d**). A third dose of S-targeting mRNA vaccine was evidenced to increase the number of RBD-specific memory B cells with expanded potency and breadth, thus contributing to the additional protection against VOCs including Omicron^14,15^, highlighting the rationale of RBD-targeting mRNA vaccine as booster. Given that more and more Chinese population have received three-dose inactivated vaccines, additional clinical trials are being conducted to assay the benefits of heterologous boosting with AWcorna.

Finally, despite the vaccine effectiveness of AWcorna booster in preventing infection by SARS-CoV-2 and other VOCs remains to be determined, the induction of potent neutralization antibodies against WT and VOCs, as well as the affordable safety profile, support the emergency use of AWcorna as heterologous booster in China. A more potent mRNA vaccine and improved booster strategy should be warranted to meet the urgent and huge need to stop the ongoing Omicron outbreaks in China and COVID-19 pandemic worldwide.

## METHODS AND MATERIALS

### Study design

We conducted a randomized clinical trial involving 300 adults (≥18 years of age) who were tested negative by RT-PCR screening for COVID-19 at the time of participation to elucidate the immunogenicity and safety of an mRNA-based vaccine (AWcorna) as a booster compared to that of homologous booster using an inactivated viral vaccine (CoronaVac).

### Ethics statement

The trial was reviewed and approved by the Research Ethics Committee of the Center for Disease Control and Prevention of Yunnan province. The study protocol and related materials were approved by the independent Ethics Committees as well, and this trial was conducted in accordance with the Declaration of Helsinki and Good Clinical Practice with the register number of ChiCTR2100053701 (NO. 2021-15). Written informed consents were obtained from each participant before the screening.

### Participants

Eligible participants met all inclusion criteria and did not trigger any exclusion criteria. Those aged 18 years and above, received full 2-dose inactivated viral priming around 6 months ago. Of the 300 participants, 175 received 2-dose CoronaVac, 14 received 2-dose BBIBP CorV, and 111 received one-dose CoronaVac and 1-dose BBIBP CorV. Participants with a previous clinical or virologic COVID-19 diagnosis or SARS-CoV-2 infection or women with positive urine pregnancy test results were excluded from this study. Participants with a medical history of convulsion, serious acute hypersensitive reaction to vaccines, acute febrile diseases or infectious diseases, congenital or acquired angioedema, asplenia or functional asplenia, thrombocytopenia or other coagulation disorders, anti-allergy therapy, or blood products within 3 months were also excluded.

### Randomization

Each participant was assigned a unique subject ID by authorized assigners successively according to a Prespecified allocation kit, which was generated by an independent randomization statistician from Beijing Key Tech Statistical Consulting Co., Ltd. via SAS software (SAS® Institute, Cary, North Carolina, USA) with the ratio of 2:1 to the AWcorna and CoronaVac groups. Since the different appearances of the two kinds of vaccines, inoculators could not keep in blind when vaccines had been used. And hence, staff who were assigned to inoculate would not be involved in any other research jobs, especially for subjects’ safety follow-up procedures. Participants would be masked when receiving the jab by a special curtain in the injection room to avoid the identification of he or she, and disclosure of the allocated group. Other investigators, laboratory staff, and outcome assessors were kept blinded also.

### Interventions

The AWcorna vaccine (15 μg/dose) (batch number RR202109006) is manufactured by Yuxi Walvax Biotechnology Co. Ltd. (an affiliate of Walvax Biotechnology Co., Ltd.), and supplied in pre-filled 0.5 ml syringes. The CoronaVac (Sinovac) vaccine, is an inactivated whole-virion vaccine with aluminum hydroxide as the adjuvant. Each dose of CoronaVac contains 3 μg SARS CoV-2 virion in a 0.5 ml aqueous suspension for injection with 0.45 mg/ml aluminum.

### Assessments

After injection, subjects received an in-site 30-minutes safety observation conducted by research staff to confirm if any immediate reactions occurred. Any adverse events (AEs) discovered or any relevant concomitant medications declared within 28 days after vaccination were recorded by subjects with the help of the daily cards and connections cards which were prepared and managed by the study team. For each subject, serious adverse events (SAEs), adverse events of interest (AESI), and pregnancy were collected from the enrollment till 12 months after his/her booster dose. Up to 6 ml of blood sample was collected from each participant at baseline pre-booster vaccination and on day 14 and day 28 after receiving the booster dose.

### Endpoints

The primary endpoints for safety were: 1) The incidence rates of adverse reactions/adverse events within 30 minutes, during Day 0-14 and Day 0-28 after vaccination; and 2) The incidence rates of adverse reactions/adverse events with a severity of grade 3 and above within 30 minutes and during Day 0-14 and Day 0-28 after vaccination. The primary immunogenicity endpoint was the titers of neutralizing antibody against wild type SARS-CoV-2 as measured by live virus neutralization assay 14 days post booster.

### Laboratory assays

The neutralizing antibodies in sera against the wild-type strain (GenBank: MT123291), Delta variant (IQTC-IM2175251), and Omicron variant (IQTC-Y216017) (Guangzhou Customs Technology Center, Guangzhou, China) were determined by using a cytopathic effect (CPE)-based microneutralization assay. Two-fold serial dilutions (starting from 1:4) of heat-inactivated sera were tested in duplicate wells for the presence of neutralizing antibodies in the monolayer of Vero E6 cells. 100 TCID50 of virus in 50 μl/well was incubated with 50 μl of serum in 96-well plates for 2 h. Vero E6 cells were trypsinized and resuspended in Dulbecco’s Modified Eagle Medium (DMEM) containing 4% of fetal bovine serum and 1% of pen/strep at a concentration of 1.2×10^5^ cells/ml and 100 μl of cells suspension were then added into the 96-well plates, followed by incubation at 37 °C, 5% CO2 for 4 days. The neutralization was determined by the appearance of CPE in images captured with Celigo Image Cytometer on day 4 post-infection. The neutralizing antibody titer was defined as the reciprocal of the highest sample dilution that protected at least 50% of cells from CPE.

RBD-specific ELISA antibody responses were measured using an indirect ELISA assay with a cutoff titer of 1:10. The commercial Anti-SARS-CoV-2 RBD IgG ELISA kit was used for detection. Measurement was performed using a Multiskan GO reader (Thermo Fisher) to detect optical density at 450 and 630 nm using SkanIt Software for Microplate Readers (version 4.1.0.43).

The WHO international standard for anti-SARS-CoV-2 IgG (NIBSC code 20/136) was used as a reference with the serum samples measured in this study for calibration of the serological assays. The WHO reference (NIBSC code: 20/136) is equivalent to a live viral neutralizing antibody titer of 1:139 against wild-type SARS-CoV-2 and a titer of 1:213 against the Delta variant B.1.617.2, while the WHO reference (1,000 BAU/ml in serum) is equivalent to an RBD-specific IgG ELISA antibody titer of 1:5,490. Live viral neutralizing antibodies against wild-type strain and the Delta variant and levels of RBD-binding IgG isotypes in serum were measured on days 0, 14, and 28 after the booster. Live viral neutralizing antibodies against the Omicron variant BA.1.1 were detected only on day 28 after the booster in a subgroup randomly selected from both groups.

### Sample size

The sample size was determined based on the hypothesis that the booster vaccination of mRNA vaccine following the two-dose inactivated vaccine regimen be non-inferior to that of the booster of inactivated vaccine in neutralizing antibody. It was assumed that the pooled standard deviation of log10-transformed neutralizing data was 0.5 and equal GMT in both the mRNA vaccine group and CoronaVac group. 200 participants in the mRNA group and 100 participants in the CoronaVac group could have at least 80% power to observe that the lower limit of the 95% confidence interval of GMT ratio between the two groups was greater than the non-inferiority margin (2/3), with the one-sided significance level of 2.5%.

### Statistical analysis

The geometric mean titer (GMT) and 95% confidence interval (CI) were used to describe neutralizing results in the mRNA vaccine group and CoronaVac group after booster vaccination, and the GMT ratio between the two groups and 95% CI were estimated. The non-inferiority result would be concluded if the lower bound of 95%CI was larger than 2/3. When the non-inferiority conclusion was concluded, the superiority would be considered sequentially if the lower bound was larger than 1. The seroconversion rate and Clopper-Pearson 95%CI after booster vaccination were estimated as well, and the difference between the two groups was calculated using the Miettinen-Nurminen method.

We assessed the number and proportion of participants with adverse reactions 0-28 days after the booster dose. For fever, besides the NMPA standard, we also derived the oral temperature by adding 0.2°C to the collected auxiliary temperature, and then re-graded the adverse reaction based on the FDA standard to provide more comparable results with marketed vaccines. We used the χ2 test or Fisher’s exact test to analyze categorical data, the t-test to analyze the log-transformed antibody titers, and the Wilcoxon rank-sum test for data not following a normal distribution. The correlation between concentrations of log-transformed neutralizing antibody and binding antibody levels was analyzed using Pearson’s correlation. The primary analysis was performed based on the per-protocol population. Statistical analyses were performed using SAS (version 9.4).

## Data Availability

All data produced in the present study are available upon reasonable request to the authors.

## ACKNOWLEDGMENTS

We thank all the participants in this trial and all staff in the research sites at Lancang CDC and Yunnan CDC. The authors acknowledge members from the Guangzhou Institute of Respiratory Health, Guangzhou Customs Technology Center, the Vazyme Medical Technology, the Beijing Key Tech Statistical Consulting, and the Beijing Stem Technology for technical support and collaboration. We sincerely thank Prof. Xuanyi Wang and Prof. Haifeng Li for critical reading and insightful suggestion for this manuscript. This work was supported in part by grant from the National Key Research and Development Program of China (2021YFC2302400) and the National Science Foundation of China (82151222). C.-F.Q. was supported by the National Science Fund for Distinguished Young Scholars (81925025), the Innovative Research Group (81621005) from the National Science Foundation of China and the Innovation Fund for Medical Sciences (2019-I2M-5-049) from the Chinese Academy of Medical Science.

## AUTHOR CONTRIBUTIONS

J.Z., Z.H. and C.-F.Q. conceived and supervised the project. X.L., J.Z., Z.W. and L.Y. designed and coordinated the experiments. J.Z. and Z.W. performed the laboratory assays. Y.L., W.H., and S.C. completed the quality assurance of this product and provided essential guidance on laboratory assays. J.S. and B.Y. provided technical expertise on mRNA vaccine production. Y.Z. and J.C. analyzed the data. L.Y., J.C., Y.-J.H., Z.X. and C.-F.Q. drafted the manuscript. All authors revised and approved the final version.

---

**Fig.S1.**
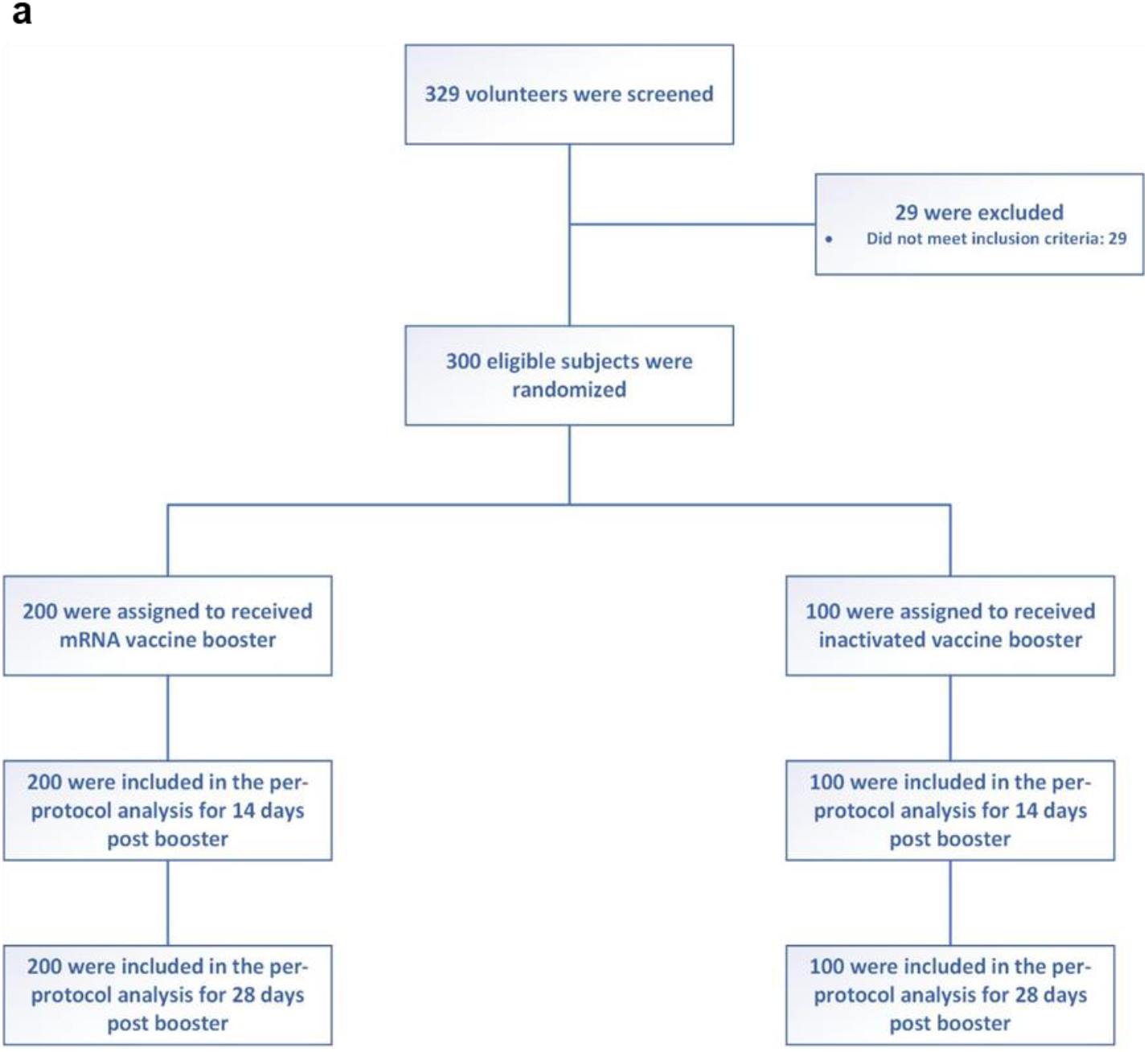
Consolidated standards of reporting trials (CONSORT) flow diagram. **a** Between the trial screening and randomization, 29 volunteers were excluded. All of them triggered the exclusion criteria. A total of 300 eligible subjects, who had received 2 doses of inactivated vaccine about 6 months ago, were randomly assigned to either the AWcorna (n=200; heterologous) or CoronaVac (n=100; homologous) booster group. All the randomized participants received vaccines at their free will and received all the scheduled follow-ups.

**Fig.S2.**
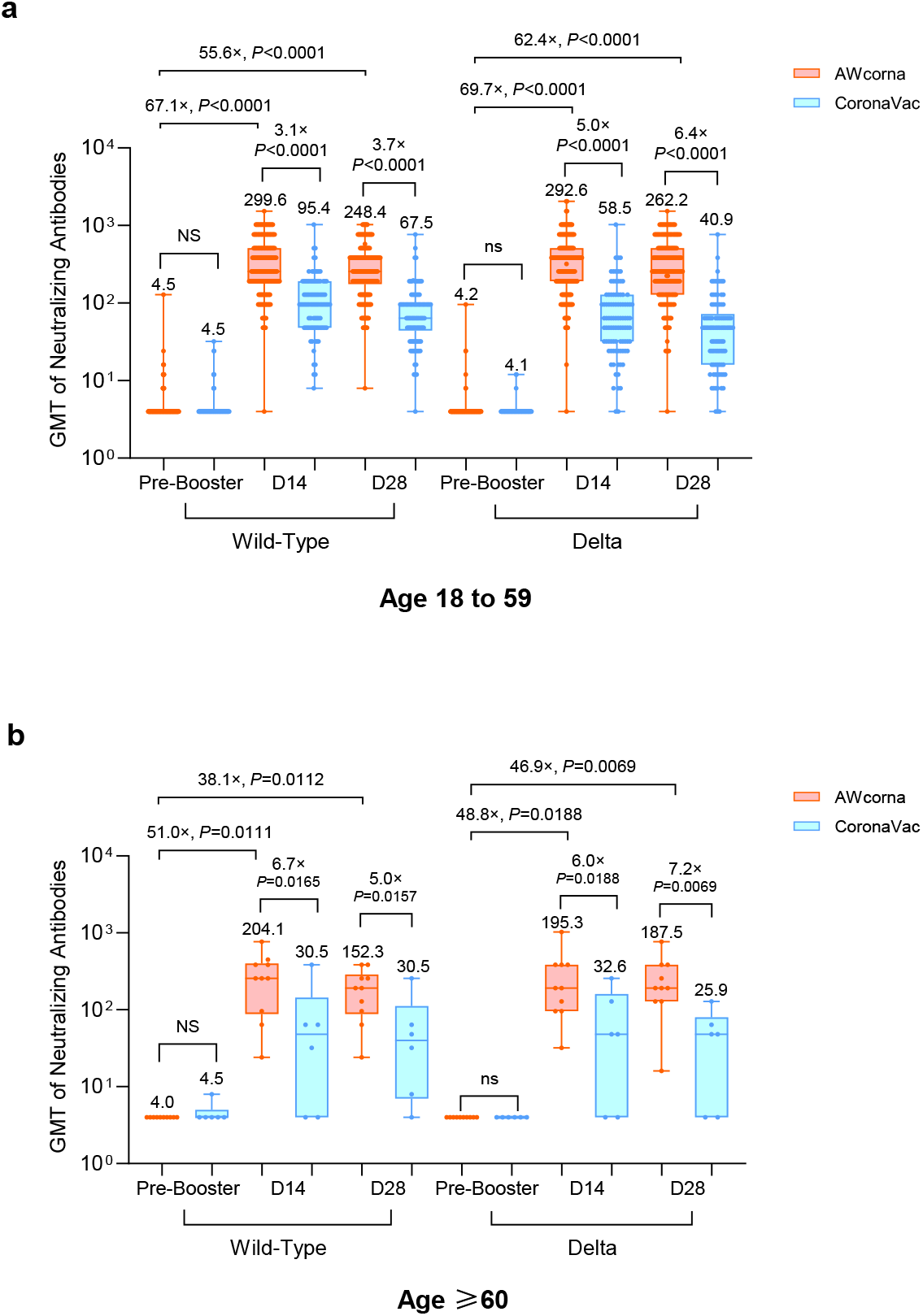
Cross neutralization against WT and Delta viruses in different age groups. **a**,**b** GMT of neutralizing antibodies to wild-type SARS-CoV-2 and Delta variant for younger adult population aged 18-59 (AWcorna=190, CoronaVac=90) (**a**) and elder population aged 60 and above (AWcorna=10, CoronaVac=6) (**b**). GMT data are presented in box-and-whisker plots. The whiskers indicate the range, the top and bottom of the boxes indicate the interquartile range, and the horizontal line within each box indicates the median. Figures with the x suffix indicate the GMT ratio between groups or GMT increase fold raised from pre-to post-booster level. *P* values were obtained from comparisons between the two treatment groups using *t*-tests for log-transformed antibody.

**Table S1.** Baseline characteristics of the trial participants, who had received two doses of priming inactivated vaccine.^*^

|  | **AWcorona** | **CoronaVac** | **P-Value** |
| --- | --- | --- | --- |
|  | **(N=200)** | **(N=100)** |  |
| **Age(Median, IQR)** | 43.0 (36.5,49.0) | 40.0 (34.0,48.5) | 0.5165 |
| **Sex, male(%)** | 116 (58.00) | 55 (55.00) | 0.6208 |
| **BMI(min-max) †** | 23.56<br>(15.58-34.53) | 23.22<br>(18.31-33.31) | 0.3998 |
| **Ethnicity‡** |  |  |  |
| Lahu, n(%) | 179 (89.50) | 88 (88.00) | 0.5418 |
| Wa, n(%) | 1 (0.50) | 0 (0.00) |  |
| Hani, n(%) | 10 (5.00) | 3 (3.00) |  |
| Yi, n(%) | 2 (1.00) | 1 (1.00) |  |
| Dai, n(%) | 0 (0.00) | 0 (0.00) |  |
| Other, n(%) | 8 (4.00) | 8 (8.00) |  |
| **RT-PCR, negative(%)** | 200(100) | 100(100) | 1.0000 |
| **Vital Signs** |  |  |  |
| Systolic Pressure | 118(81-141) | 118(85-140) | 0.9615 |
| Diastolic Pressure | 73(48- 89) | 74(56-90) | 0.3679 |
| Pulse | 79(56-114) | 81(53-110) | 0.2182 |
| **Comorbidities** |  |  |  |
| **Any (%)** | 9 | 9 | 1.0000 |
| Gastrointestinal disorders (%) | 3.5 | 2 | 0.7227 |
| Vascular disorders (%) | 1 | 2 | 0.6030 |
| Respiratory, thoracic and mediastinal disorders (%) | 1.5 | 0 | 0.5533 |
| Infections and infestations (%) | 1 | 1 | 1.0000 |
| Musculoskeletal and connective tissue disorders (%) | 0.5 | 1 | 1.0000 |
| Nervous system disorders (%) | 0.5 | 1 | 1.0000 |
| Blood and lymphatic system disorders (%) | 1 | 0 | 0.5541 |
| Endocrine disorders (%) | 0 | 1 | 0.3333 |
| Investigations (%) | 0.5 | 0 | 1.0000 |
| Reproductive system and breast disorders (%) | 0 | 1 | 0.3333 |
| **SARS-CoV-2 specific antibody titers on day 0§** |  |  |  |
| Geometric Mean Titer of anti-RBD IgG (Mean, 95%CI) | 134.7 | 140.8 | 0.7061 |
|  | (118.3, 153.4) | (115.3, 172.0) |  |
| Geometric Mean Titer of nAb against Wild-Type SARS-CoV-2 (Mean, 95%CI) |  |  |  |
|  | 4.4(4.2, 4.7) | 4.5(4.2, 4.9) | 0.7831 |
| Geometric Mean Titer of nAb against Delta Variant (Mean, 95%CI) |  |  |  |
|  | 4.2(4.0, 4.4) | 4.1(4.0, 4.3) | 0.5951 |

\* The first trial visit occurred before the booster, and the second and third trial visit occurred 14- and 28-days after administration of the booster. IQR denotes interquartile range.
† The body-mass index (BMI) is the weight in kilograms divided by the square of the height in meters.
‡ Categories were reported by the participants. The categories shown are those used by the investigators to denote ethnicity.
§ In the two groups, blood was drawn on day 0, which was the day of the first trial visit. Day 0 was also the day of the booster in the two groups.

**Table S2.**
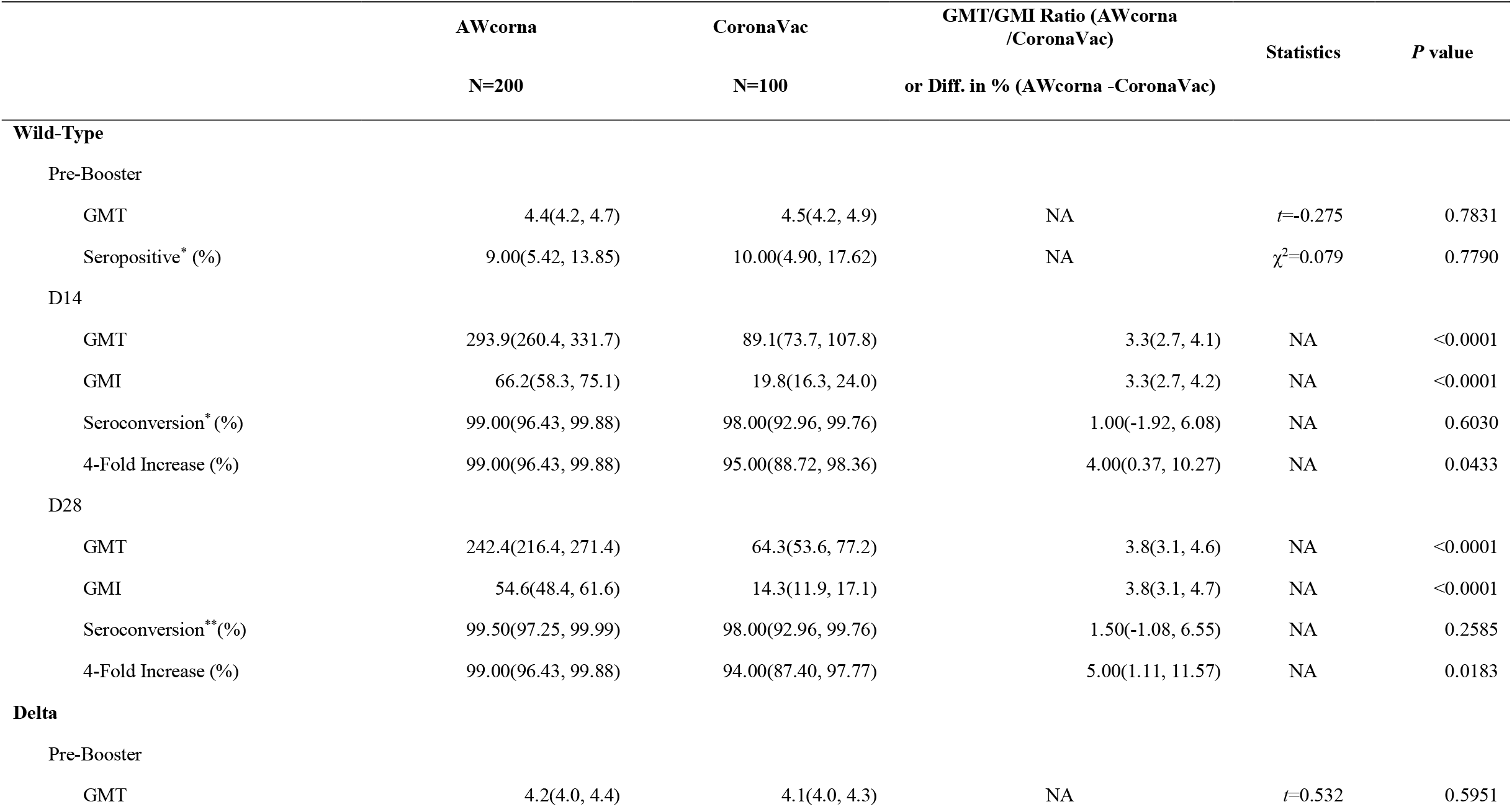
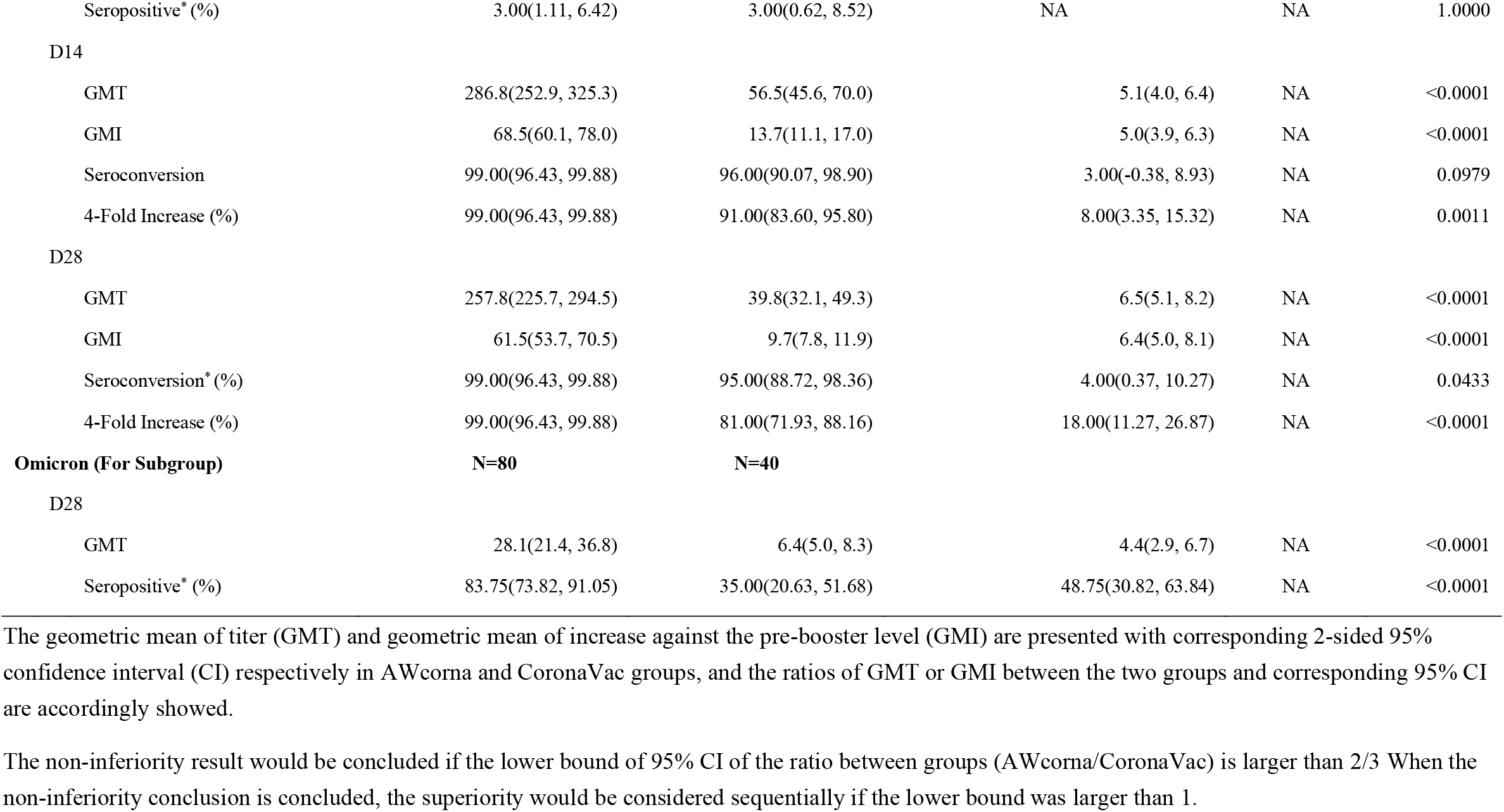
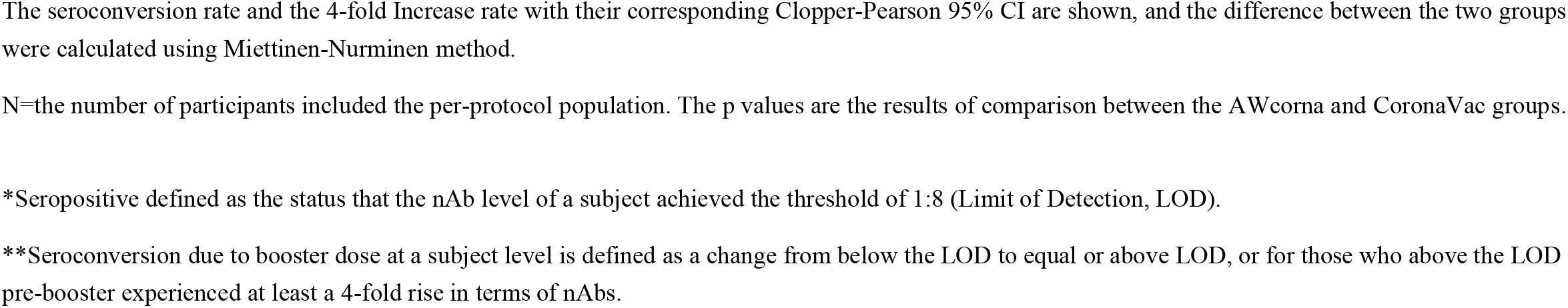
Neutralizing antibodies to the wild-type SARS-CoV-2 as well as Delta and Omicron variants before and after booster.

**Table S3.**
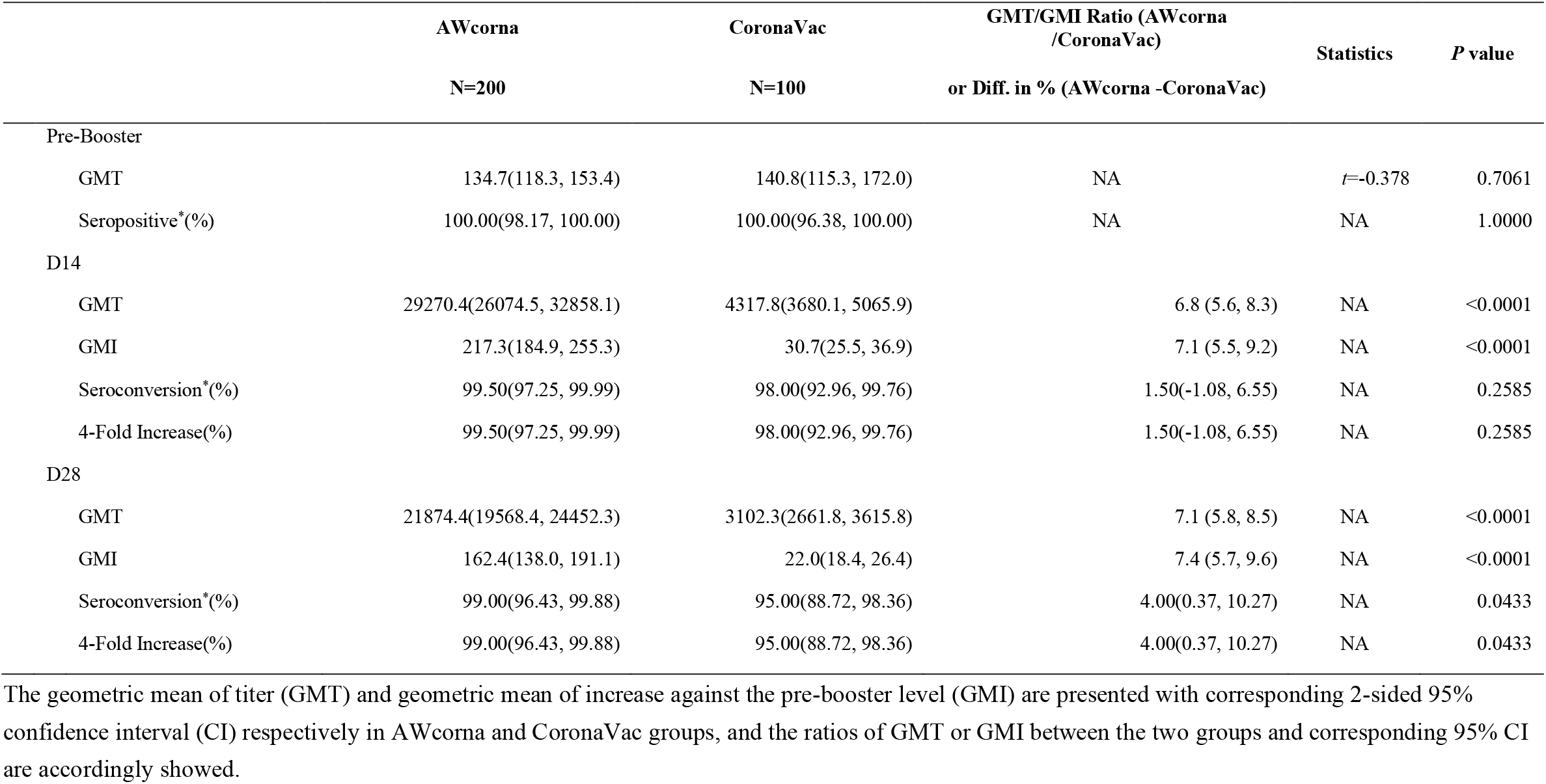
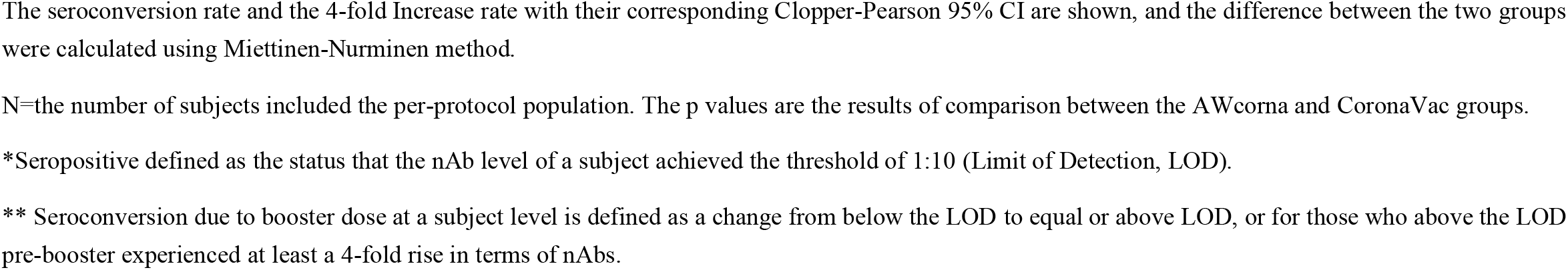
RBD-Specific antibodies before and after booster.

**Table S4.** Solicited and unsolicited adverse reactions that occurred within 28 days after booster vaccination.

| Event Name | AWcorna<br>(N=200) | CoronaVac (N=100) | P-Value |
| --- | --- | --- | --- |
| Severity | n(%) | n(%) |  |
| **AE (Any)** | 139(69.5) | 20(20) | **<0.0001** |
| Grade1 | 85(42.5) | 12(12) | **<0.0001** |
| Grade2 | 71(35.5) | 6(6) | **<0.0001** |
| Grade3 | 22(11) | 3(3) | **0.0246** |
| Grade4 | 0(0) | 0(0) | 1 |
| Grade5 | 0(0) | 0(0) | 1 |
| ≥Grade2 | 84(42) | 9(9) | **<0.0001** |
| ≥Grade3 | 22(11) | 3(3) | **0.0246** |
| **Solicited AE (Any)** | 138(69) | 20(20) | **<0.0001** |
| Grade1 | 85(42.5) | 12(12) | **<0.0001** |
| Grade2 | 69(34.5) | 6(6) | **<0.0001** |
| Grade3 | 22(11) | 3(3) | **0.0246** |
| Grade4 | 0(0) | 0(0) | 1 |
| ≥Grade2 | 83(41.5) | 9(9) | **<0.0001** |
| ≥Grade3 | 22(11) | 3(3) | **0.0246** |
| **Systemic (Any)** | 131(65.5) | 20(20) | **<0.0001** |
| Grade1 | 73(36.5) | 12(12) | **<0.0001** |
| Grade2 | 67(33.5) | 6(6) | **<0.0001** |
| Grade3 | 20(10) | 3(3) | **0.0372** |
| Grade4 | 0(0) | 0(0) | 1 |
| ≥Grade2 | 82(41) | 9(9) | **<0.0001** |
| ≥Grade3 | 20(10) | 3(3) | **0.0372** |
| **Fever (FDA Standard, Any)** | 67(33.5) | 4(4) | **<0.0001** |
| Grade1 | 39(19.5) | 2(2) | **<0.0001** |
| Grade2 | 20(10) | 2(2) | **0.0100** |
| Grade3 | 8(4) | 0(0) | 0.0555 |
| Grade4 | 0(0) | 0(0) | 1 |
| ≥Grade2 | 28(14) | 2(2) | **0.0008** |
| ≥Grade3 | 8(4) | 0(0) | 0.0555 |
| Diarrhea (Any) | 0(0) | 0(0) | 1 |
| Grade1 | 0(0) | 0(0) | 1 |
| Grade2 | 0(0) | 0(0) | 1 |
| Grade3 | 0(0) | 0(0) | 1 |
| Grade4 | 0(0) | 0(0) | 1 |
| ≥Grade2 | 0(0) | 0(0) | 1 |
| ≥Grade3 | 0(0) | 0(0) | 1 |
| Nausea (Any) | 5(2.5) | 1(1) | 0.6674 |
| Grade1 | 4(2) | 0(0) | 0.3052 |
| Grade2 | 1(0.5) | 0(0) | 1 |
| Grade3 | 0(0) | 1(1) | 0.3333 |
| Grade4 | 0(0) | 0(0) | 1 |
| ≥Grade2 | 1(0.5) | 1(1) | 1 |
| ≥Grade3 | 0(0) | 1(1) | 0.3333 |
| Vomiting (Any) | 5(2.5) | 1(1) | 0.6674 |
| Grade1 | 5(2.5) | 0(0) | 0.1734 |
| Grade2 | 0(0) | 0(0) | 1 |
| Grade3 | 0(0) | 1(1) | 0.3333 |
| Grade4 | 0(0) | 0(0) | 1 |
| ≥Grade2 | 0(0) | 1(1) | 0.3333 |
| ≥Grade3 | 0(0) | 1(1) | 0.3333 |
| Headache (Any) | 52(26) | 7(7) | **<0.0001** |
| Grade1 | 11(5.5) | 3(3) | 0.3993 |
| Grade2 | 41(20.5) | 3(3) | **<0.0001** |
| Grade3 | 0(0) | 1(1) | 0.3333 |
| Grade4 | 0(0) | 0(0) | 1 |
| ≥Grade2 | 41(20.5) | 4(4) | **<0.0001** |
| ≥Grade3 | 0(0) | 1(1) | 0.3333 |
| Muscle Aches (Any) | 15(7.5) | 1(1) | **0.0255** |
| Grade1 | 5(2.5) | 0(0) | 0.1734 |
| Grade2 | 9(4.5) | 1(1) | 0.1732 |
| Grade3 | 1(0.5) | 0(0) | 1 |
| Grade4 | 0(0) | 0(0) | 1 |
| ≥Grade2 | 10(5) | 1(1) | 0.1073 |
| ≥Grade3 | 1(0.5) | 0(0) | 1 |
| Joint Pain (Any) | 6(3) | 1(1) | 0.4310 |
| Grade1 | 3(1.5) | 0(0) | 0.5533 |
| Grade2 | 2(1) | 1(1) | 1 |
| Grade3 | 1(0.5) | 0(0) | 1 |
| Grade4 | 0(0) | 0(0) | 1 |
| ≥Grade2 | 3(1.5) | 1(1) | 1 |
| ≥Grade3 | 1(0.5) | 0(0) | 1 |
| Chills (Any) | 10(5) | 1(1) | 0.1073 |
| Grade1 | 7(3.5) | 1(1) | 0.2765 |
| Grade2 | 3(1.5) | 0(0) | 0.5533 |
| Grade3 | 0(0) | 0(0) | 1 |
| Grade4 | 0(0) | 0(0) | 1 |
| ≥Grade2 | 3(1.5) | 0(0) | 0.5533 |
| ≥Grade3 | 0(0) | 0(0) | 1 |
| Fatigue (Any) | 4(2) | 0(0) | 0.3052 |
| Grade1 | 2(1) | 0(0) | 0.5541 |
| Grade2 | 1(0.5) | 0(0) | 1 |
| Grade3 | 1(0.5) | 0(0) | 1 |
| Grade4 | 0(0) | 0(0) | 1 |
| ≥Grade2 | 2(1) | 0(0) | 0.5541 |
| ≥Grade3 | 1(0.5) | 0(0) | 1 |
| Rash (Not at Injection Site, Any) | 0(0) | 1(1) | 0.3333 |
| Grade1 | 0(0) | 0(0) | 1 |
| Grade2 | 0(0) | 0(0) | 1 |
| Grade3 | 0(0) | 1(1) | 0.3333 |
| Grade4 | 0(0) | 0(0) | 1 |
| ≥Grade2 | 0(0) | 1(1) | 0.3333 |
| ≥Grade3 | 0(0) | 1(1) | 0.3333 |
| Hypersensitivity (Any) | 1(0.5) | 0(0) | 1 |
| Grade1 | 0(0) | 0(0) | 1 |
| Grade2 | 1(0.5) | 0(0) | 1 |
| Grade3 | 0(0) | 0(0) | 1 |
| Grade4 | 0(0) | 0(0) | 1 |
| ≥Grade2 | 1(0.5) | 0(0) | 1 |
| ≥Grade3 | 0(0) | 0(0) | 1 |
| Local AE (Any) | 34(17) | 2(2) | **<0.0001** |
| Grade1 | 26(13) | 2(2) | **0.0013** |
| Grade2 | 7(3.5) | 0(0) | 0.0998 |
| Grade3 | 2(1) | 0(0) | 0.5541 |
| Grade4 | 0(0) | 0(0) | 1 |
| ≥Grade2 | 9(4.5) | 0(0) | **0.0319** |
| ≥Grade3 | 2(1) | 0(0) | 0.5541 |
| Pain at Injection Site (Any) | 34(17) | 2(2) | **<0.0001** |
| Grade1 | 25(12.5) | 2(2) | **0.0021** |
| Grade2 | 7(3.5) | 0(0) | 0.0998 |
| Grade3 | 2(1) | 0(0) | 0.5541 |
| Grade4 | 0(0) | 0(0) | 1 |
| ≥Grade2 | 9(4.5) | 0(0) | **0.0319** |
| ≥Grade3 | 2(1) | 0(0) | 0.5541 |
| Induration (Any) | 0(0) | 0(0) | 1 |
| Grade1 | 0(0) | 0(0) | 1 |
| Grade2 | 0(0) | 0(0) | 1 |
| Grade3 | 0(0) | 0(0) | 1 |
| Grade4 | 0(0) | 0(0) | 1 |
| ≥Grade2 | 0(0) | 0(0) | 1 |
| ≥Grade3 | 0(0) | 0(0) | 1 |
| Redness (Any) | 0(0) | 0(0) | 1 |
| Grade1 | 0(0) | 0(0) | 1 |
| Grade2 | 0(0) | 0(0) | 1 |
| Grade3 | 0(0) | 0(0) | 1 |
| Grade4 | 0(0) | 0(0) | 1 |
| ≥Grade2 | 0(0) | 0(0) | 1 |
| ≥Grade3 | 0(0) | 0(0) | 1 |
| Rash (at Injection Site, Any) | 0(0) | 0(0) | 1 |
| Grade1 | 0(0) | 0(0) | 1 |
| Grade2 | 0(0) | 0(0) | 1 |
| Grade3 | 0(0) | 0(0) | 1 |
| Grade4 | 0(0) | 0(0) | 1 |
| ≥Grade2 | 0(0) | 0(0) | 1 |
| ≥Grade3 | 0(0) | 0(0) | 1 |
| Swelling (Any) | 2(1) | 0(0) | 0.5541 |
| Grade1 | 2(1) | 0(0) | 0.5541 |
| Grade2 | 0(0) | 0(0) | 1 |
| Grade3 | 0(0) | 0(0) | 1 |
| Grade4 | 0(0) | 0(0) | 1 |
| ≥Grade2 | 0(0) | 0(0) | 1 |
| ≥Grade3 | 0(0) | 0(0) | 1 |
| Itch (Any) | 0(0) | 0(0) | 1 |
| Grade1 | 0(0) | 0(0) | 1 |
| Grade2 | 0(0) | 0(0) | 1 |
| Grade3 | 0(0) | 0(0) | 1 |
| Grade4 | 0(0) | 0(0) | 1 |
| ≥Grade2 | 0(0) | 0(0) | 1 |
| ≥Grade3 | 0(0) | 0(0) | 1 |
| Unsolicited AE (Any) | 2(1) | 1(1) | 1 |
| Grade1 | 0(0) | 0(0) | 1 |
| Grade2 | 2(1) | 0(0) | 0.5541 |
| Grade3 | 0(0) | 1(1) | 0.3333 |
| Grade4 | 0(0) | 0(0) | 1 |
| Grade5 | 0(0) | 0(0) | 1 |
| ≥Grade2 | 2(1) | 1(1) | 1 |
| ≥Grade3 | 0(0) | 1(1) | 0.3333 |

Data are n (%). n, number of participants; %, percentage of participants; any, all participants with any grade of adverse reactions. The analysis was based on the per-protocol population. *P* values shown in bold are <0.05.

